# MOG1^L18F^-mediated increase in late sodium current produces Long QT Syndrome

**DOI:** 10.64898/2025.12.12.25340695

**Authors:** Paula G. Socuéllamos, Juan Manuel Ruiz-Robles, Francisco M. Cruz, Álvaro Macías, Alba Vera-Zambrano, Ana I Moreno-Manuel, Carmen Prior, Antonio J. Cartón, Carmen Valenzuela, José Jalife

## Abstract

**Background:** Na_V_1.5 channels, encoded by *SCN5A*, are essential for the genesis and shaping of the cardiac action potential (AP). *Gain-of-function* (GoF) variants in *SCN5A* are associated with long QT syndrome (LQTS), whereas loss-of-function (LoF) mutations are linked with Brugada syndrome. MOG1 is an integral part of the Na_V_1.5 channelosome, increasing both current and membrane expression of Na_V_1.5. Two LoF variants in MOG1 (E61X and E83D) cause Brugada Syndrome in patients, but no association with LQTS has been reported.

**Methods:** We identified the first variant of unknown significance (VUS) in MOG1 (g.52C>T; p.18L>F) in a proband with LQTS. We generated AAV9-mediated cardiac-specific mouse models expressing MOG1^WT^ and MOG1^L18F^. We performed surface ECG, programmed electrical stimulation in live mice, optical mapping in intact hearts, and patch-clamping, Ca^2+^ dynamics and molecular biology assays in ventricular cardiomyocytes and HEK293 cells.

**Results:** Clinical data from the MOG1^L18F^ proband revealed complete AV block, prolonged QT intervals, and non-sustained ventricular tachycardia (nsVT) under hypokalemia. MOG1^L18F^ mice recapitulated the patient’s phenotype, with QT prolongation and an increased incidence of arrhythmia, which worsened upon hypokalemia. Voltage-clamp recordings revealed a marked increase in the late sodium current (*I*_NaL_) in MOG1^L18F^, accompanied by Na_V_1.8 expression enhancement in the sarcolemma. In current-clamp and optical mapping experiments, action potential duration (APD) increased dramatically at low stimulation frequencies resulting in a very steep APD restitution curve in MOG1^L18F^ ventricular cardiomyocytes. This was associated with a high incidence of early and delayed afterdepolarizations (EADs and DADs, respectively) and triggered activity. Notably, the cellular electrophysiological effects of MOG1^L18F^ were reversed by the Na_V_1.8 inhibitor A-803467, which also abridged the QT prolongation and reduced the arrhythmia inducibility in MOG1^L18F^ mice, with no effect in MOG1^WT^ mice.

**Conclusion:** We have uncovered a new genetic basis for LQTS. Our findings demonstrate that the MOG1^L18F^ variant impairs AV conduction, increases I_NaL_, opposes repolarizing currents, and prolongs both action potential duration and the QT interval, ultimately leading to ventricular arrhythmias, particularly under hypokalemic conditions. As Na_V_1.8 mediates the pathogenic I_NaL_ increase and interacts with both MOG1 and Na_V_1.5, it emerges as a promising therapeutic target.

## INTRODUCTION

Na_V_1.5 channels, encoded by *SCN5A*, are essential for the genesis of the cardiac action potential (AP), and their late current, *I*_NaL_, contributes to its duration^1,2^. MOG1, encoded by *RANGRF*, is a chaperone that interacts with Na_V_1.5 through its cytoplasmic loops I^3^ and II^4,5^, favoring channel trafficking to the plasma membrane and increasing *I*_Na_ without modifying the biophysical properties of the channel^3,4,6^. MOG1 is primarily expressed in the heart, and even though according to ClinVar, 178 variants in *RANGRF* have been found in patients with cardiac arrhythmia (**Extended table**), only two MOG1 gene variants (E61X and E83D) have been functionally characterized^7–9^. MOG1^E61X^ and MOG1^E83D^ are associated with Brugada syndrome, both reducing trafficking to the sarcolemma and Na_V_1.5 current amplitude^7–9^. MOG1^L18F^ was found in two homozygous twins with hypertrophic cardiomyopathy, whose pathogenic mechanisms remain unexplored^10^. *Loss-of-function* mutations in *SCN5A* produce Brugada syndrome, dilated cardiomyopathy, and sick sinus syndrome, which can be rescued by MOG1^11,12^, thus demonstrating the relevance of this protein in cardiac electrophysiology. On the other hand, *gain-of-function* variants in *SCN5A* are associated with type 3 Long QT syndrome (LQTS; ORPHA: 768) due to an increased *I*_NaL_^13–15^. LQTS is a pathophysiological condition associated with an increased risk of syncope and sudden cardiac death (SCD) due to ventricular tachyarrhythmias^16^. Even though the magnitude of *I*_NaL_ is relatively small compared with the fast *I*_Na_ in cardiomyocytes, it contributes significantly to the AP shape and duration^16^. Cardiac *I*_NaL_ is modulated by CaMKII, Na_V_β subunits, PKA and PKC phosphorylation, and non-cardiac Na_V_ channels, like Na_V_1.8, encoded by *SCN10A*^16,17^.

Recent research has focused on deciphering the expression and role of Na_V_1.8 in the heart. Apart from its expression in nociceptive sensory neurons^18^, Na_V_1.8 is also expressed in intracardiac neurons^19^, the cardiac specialized conduction system^20^, and atrial and ventricular cardiomyocytes^21–23^. *SCN10A*/Na_V_1.8 is of great clinical importance despite its low expression in the mammalian heart. *SCN10A* mutations have been found in patients with Brugada syndrome^23–25^, atrial fibrillation^26,27^, LQTS^28^, hypertrophic cardiomyopathy^29^, and SCD^30^. Its gene product is increased in heart failure, thus increasing *I*_NaL_^31,32^. Studies conducted in mouse and rabbit cardiomyocytes deciphered the involvement of Na_V_1.8 in cardiac electrophysiology^21,33,34^, and heterologous systems demonstrated that Na_V_1.8 and Na_V_1.5 physically interact and are coregulated^21,23,35^. Moreover, recent research described a short, cardiac-specific isoform of *SCN10A*/Na_V_1.8 (*SCN10A-s*/Na_V_1.8-s) —comprising the last two transmembrane segments of DIII (S5 and S6), the DIII-DIV linker, DIV, and the C-terminal—that enhances Na_V_1.5 current in a similar way as the full-length isoform^23,36^.

We report the identification and characterization of the first MOG1 variant, L18F, in a patient with long QT syndrome (LQTS). To investigate the molecular mechanisms underlying the life-threatening arrhythmias caused by the MOG1^L18F^ variant, we generated a mouse model of LQTS using adeno-associated virus (AAV)-mediated MOG1 gene transfer. Employing a multidisciplinary approach—including patch-clamping, intracardiac electrophysiological stimulation, and optical mapping—we demonstrate that MOG1^L18F^ enhances late sodium current (I_NaL_) via Na_V_1.8, resulting in a persistent depolarizing current during the AP plateau. The increased I_NaL_ opposes repolarizing currents, prolongs AP duration (APD) and the QT interval, thereby recapitulating the patient’s arrhythmogenic LQTS phenotype. This is the first evidence that a MOG1 variant can increase I_NaL_ and cause LQTS.

## MATERIAL AND METHODS (see further details in the Supplementary file)

### Clinical evaluation

A 6-10-year-old boy was admitted after an incidental finding of bradycardia when being assessed for an upper respiratory tract infection. He was evaluated with an echocardiogram (Echo), cardiac magnetic resonance (MR), and several lab tests. Informed consent from the legal tutor was obtained for the use of anonymized clinical data and images with scientific objectives and literature, in the Hospital Universitario La Paz of Madrid, Spain. The study conforms to the Declaration of Helsinki.

### Mice

C57BL/6J male mice, 4-5 weeks old, were obtained from the Charles River Laboratories, and reared and housed in accordance with CNIC animal facility guidelines and regulations.

### Power analysis

To determine the statistical power and minimum sample size in our experiments, we used a power calculation using the R base “power.t.test” or “power.anova.test” function, depending on the data analysis. We used a significance level (alpha=0.05), power (1-beta=80%), the estimated difference between control and experimental data for the t-test, and estimated variances for ANOVA.

### Adeno-associated virus vector (AAV) production, purification, and mouse model generation

AAV vectors were generated using the cardiomyocyte-specific cardiac TroponinT proximal promoter (cTnT) to encode human MOG1^WT^ or MOG1^L18F^, followed by the tdTomato reporter. Vectors were packaged into AAV serotype 9 (AVV9) and produced by the triple transfection method, using HEK293T cells as described previously^37^. Mice were anesthetized with ketamine (60 mg/kg) and xylazine (20 mg/kg) via the intraperitoneal (i.p.) route. Thereafter, 3.5×10^10^ virus particles were inoculated intravenously through the femoral vein in a final volume of 50 μL. Only well-inoculated animals were included in the studies.

### Drugs

A-803467 (Sigma) was dissolved in 5% dimethyl sulfoxide (DMSO) and 95% polyethylene glycol (PEG400) as previously described^21^. For *in vivo* experiments, the stock was 15 mM and a dose of 20 mg/kg was inoculated i.p. For *in vitro* experiments, the stock was 1 mM, being further diluted to a final concentration of 30 nM^21^. KN-93 (1 µM) was incubated for 1-2 h before patch-clamp experiments and was also perfused in the extracellular solution as previously reported^38^.

### Surface electrocardiographic (ECG) recordings

Mice were anesthetized using isoflurane inhalation (0.8-1.0% volume in oxygen), and the efficacy of the anesthesia was determined by monitoring breathing rate. Six-lead surface ECGs were recorded for 5 minutes, from subcutaneous 23-gauge needle electrodes attached to each limb and connected to an MP36R amplifier unit (BIOPAC Systems). We used AcqKnowledge 4.1 software to analyze all ECG waves, segments, and interval durations in a blind and paired manner. We applied Framingham’s formula to correct the QT interval to the heart rate^39^.

### Intracardiac recording and stimulation

An octopolar catheter (Science) was inserted through the jugular vein and advanced into the right atrium and ventricle as described previously^39–41^. Ventricular arrhythmia inducibility was assessed by applying trains of twenty pulses at frequencies between 2 and 30 Hz.

### Ex-Vivo High-resolution Optical Mapping

Optical mapping experiments were carried out as previously described^42–44^. The potentiometric dye Di-4-ANEPPS (Molecular Probes) was added to achieve a final concentration of 10 µM. We used a custom-made upright 128 × 128-pixel eVolve EMCCD camera (Photometrics) running at 1,000 frames per second. Blebbistatin (10 µM) was used to reduce contraction. Stimulation at various frequencies was induced through the apex via a custom-made bipolar electrode connected to a programmable stimulator (Cibertec). Phase, activation, and APD maps were generated utilizing a custom-made pipeline developed in Matlab. The stimulation protocol was similar to intracardiac stimulation^39^.

### Cardiomyocyte isolation

Mouse ventricular cardiomyocytes were isolated as previously described^39^.

### Cell culture and transient transfection

HEK293 and HEK293T stably expressing human cardiac sodium channel gene *SCN5A* -referred to as HEK/Na_V_1.5 cells -were grown in their corresponding supplemented medium at 37 °C in a 5% CO_2_ humidified atmosphere. Transient transfection was performed using JetPRIME (Polyplus) following the manufacturer’s instructions (see Supplementary Methods).

### Patch clamping

The whole-cell patch-clamp technique, internal and external solutions, ionic currents, action potential acquisition, and data analysis were similar to those previously described for cell lines and mouse cardiomyocytes^39,41,45,46^.

### Calcium dynamics assays

Cytosolic Ca^2+^ was monitored according to previously described protocols^39^. Briefly, cells were loaded with Fluo-4-AM (Invitrogen). Fluorescence was detected in line scan mode (2 ms/scan), with the line drawn approximately through the cell center and parallel to its long axis.

### RNA extraction and qPCR

*Scn5a, Scn10a and Scn10a-s* expression was measured in isolated cardiomyocytes as previously described^41^ and using the primers described previously^23^ to detect *Scn10a*.

### Membrane fractioning, Immunoprecipitation and Western blot

For total protein and membrane fractioning, isolated ventricular MOG1^WT^ and MOG1^L18F^ cardiomyocytes were homogenized and processed according to the manufacturer’s specifications (Abcam, ab65400). HEK293 cells were homogenized as previously described^47^. Protein was quantified using the Pierce BCA Protein Assay Kit (Thermo Fisher). We used Protein G Sepharose® beads incubated with the corresponding antibodies for coimmunoprecipitation assays. Protein extracts were separated by 4-15% SDS-PAGE gels (Mini-PROTEAN TGX, BioRad, 4568084), electrotransferred onto a 0.2-µm PVDF membrane (BioRad, 1704150), and probed with specific antibodies. Please see further details in the Supplemental Methods.

## RESULTS

### The authors declare that all supporting data are available within the paper and its supplementary Information

#### Identification of MOG1^L18F^ in a patient with LQTS and life-threatening arrhythmias

The patient was admitted after an incidental finding of bradycardia when being assessed for an upper respiratory tract infection. He had no symptoms during daily activities, and his medical history had been uneventful. His parents were healthy, with normal ECG, and maternal titters for autoantibodies (anti-rho, anti-la) were negative. The resting ECG showed a complete atrio-ventricular (AV) block pattern with an escape rhythm, and narrow QRS complexes with a prolonged QT interval (>500 ms) (**Figure 1A**). On 24-hour Holter monitoring, 2:1 AV block was present for most of the recording, with a mean ventricular rate of 45 bpm (range 29 to 102 bpm), and a maximum RR interval of 2.02 s; the QTc was markedly prolonged, reaching up to 592 ms at heart rates around 60 bpm, and showing little shortening at higher rates. Echo and cardiac MRI of the proband were normal, with no signs of inflammation, edema, or fibrosis. Laboratory analysis did not find ionic imbalances; troponin and prohormone N-terminal of the brain natriuretic peptide (NT-proBNP) were negative, and the TSH level was normal. Serologies, including testing for Lyme disease, were negative.

**Figure 1.**
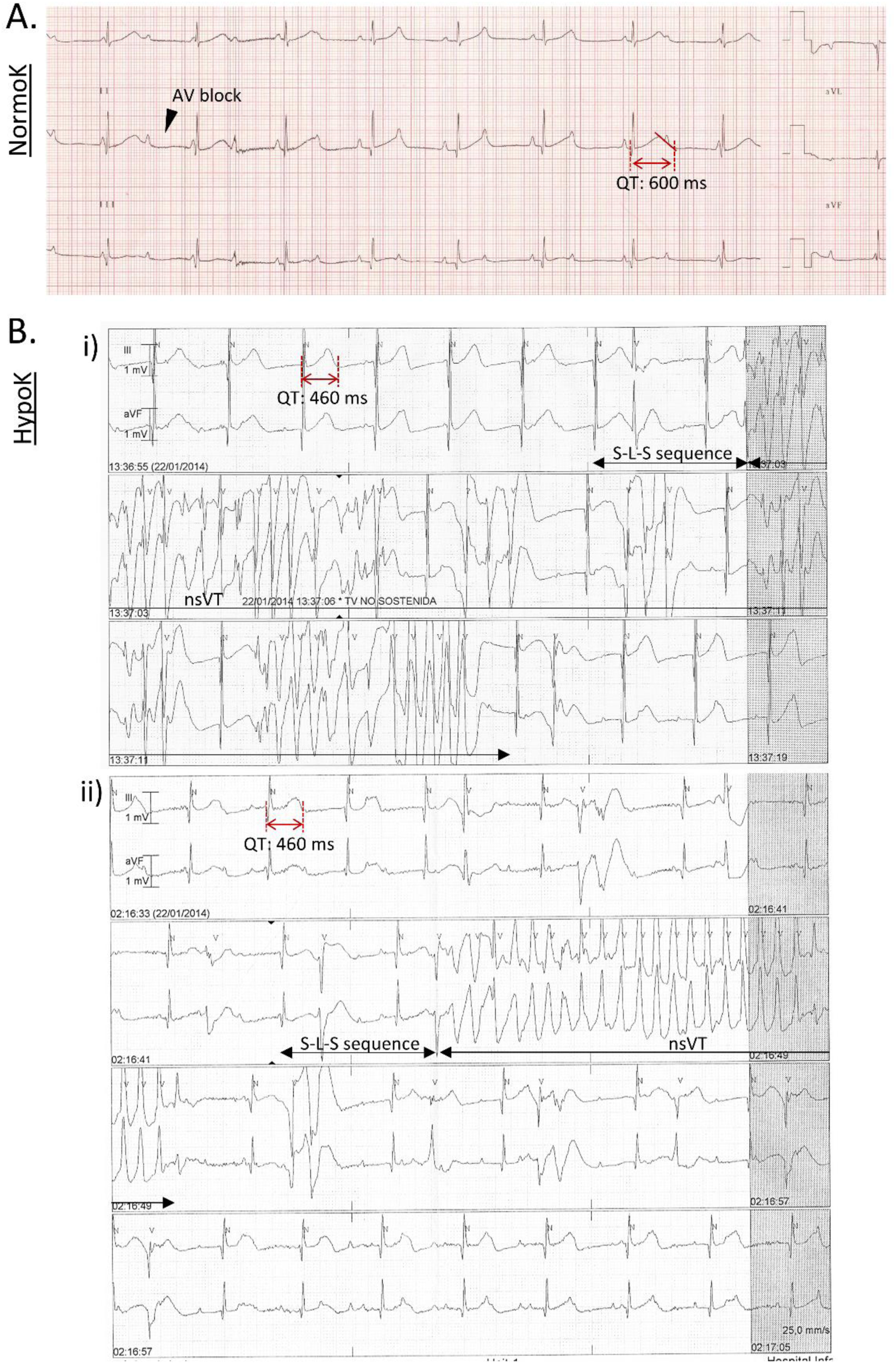
Patient’s electrical phenotype. **A.** ECG recording showing absolute QT intervals of nearly 520 ms during 2:1 AV block. **B.** Self-limited runs of non-sustained ventricular tachycardia (nsVT) on telemetry ECG monitoring during hypokalemia. **i)** First row shows polymorphic ventricular tachycardia (PVT) onset: the first beat is coupled at the end of the preceding T wave with a long interval. Previously note the triggering short-long-short (S-L-S) sequence and the unaccommodated, prolonged QT interval, both typical features of *Torsade de Pointes* PVT onset. Second and third rows show the non-sustained nature of the PVT episode, reverting to sinus rhythm with prolonged QT interval. **ii)** Second row shows PVT onset preceded by a S-L-S sequence. Third and fourth rows show the VT episode reverting to sinus rhythm with prolonged QT interval.

During admission, the patient experienced frequent vomiting that resulted in hypokalemia (serum K^+^: 2.6-2.9 mM). On telemtetry ECG, self-limited runs of non-sustained ventricular tachycardia (nsVT) were detected (**Figure 1B**), with features compatible with polymorphic VT in the form of *Torsades de pointes* (**Figure 1Bi**). Runs were preceded by a short-long-short (S-L-S) sequence, and the coupling interval of the first VT beat was 440 ms (**Figure 1Bi-ii**). The QT interval was only mildly increased at that time (450-460 ms) (**Figure 1Bi-ii**). The patient was transferred to the intensive care unit. Isoproterenol infusion and correction of ionic imbalance prevented recurrence of the VT. Ultimately, a permanent pacemaker was indicated and implanted, and nadolol was started afterwards, with good tolerance. Evolution was uneventful, and no VT recurrence was detected. Due to the similarity of the patient’s phenotype to LQT3, a genetic analysis directed towards *SCN5A* was performed, obtaining no variants in that gene. Subsequently, genetic analysis with a panel of cardiac arrhythmia-related genes was performed in the patient’s sample. This analysis revealed a single-base substitution at nucleotide 52 (g.52C>T) in the *RANGRF* gene, resulting in a change from leucine to phenylalanine (p.18L>F) in the MOG1 protein, with unknown significance and a minor allele frequency (MAF) of 0.0006 %. Variants L18F and L18I in MOG1 had been identified in patients with cardiac arrhythmias in ClinVar and Varsome, but no functional characterization was available (**Extended table**). This variant was selected due to the described interaction of MOG1 with Na_V_1.5^6^.

#### MOG1^L18F^ mice exhibit LQTS, cardiac conduction defects and arrhythmias

To characterize the functional implications of the L18F variant in MOG1, we generated AAV9-mediated cardiac-specific mouse models expressing MOG1^WT^ and MOG1^L18F^. Similarly to the LQTS patient, no structural changes or fibrosis were found in MOG1^L18F^ hearts. Histology (H&E and Masson trichome staining) (**Figure S1A**), heart-to-body weight (HW/BW) ratio determination (**Figure S1B**), and cardiomyocyte size showed no differences in MOG1^L18F^ versus MOG1^WT^ mice (**Figure S1C**). In addition, total protein analysis demonstrated that isolated mouse cardiomyocytes expressing MOG1^L18F^ and MOG1^WT^ had similar MOG1 protein expression levels (**Figure S1D**).

On surface ECG, MOG1^L18F^ mice recapitulated the patient’s phenotype, exhibiting QT and QTc (corrected by Framingham’s formula) prolongation (**Figure 2A and B**) in comparison to MOG1^WT^ animals. Other ECG parameters, including QRS and PR intervals, were not altered. Additionally, the QT interval of MOG1^L18F^ mice had a greater dependence on the RR interval, with a steeper slope (s) (**Figure 2B**). On catheter-based intracardiac programmed electrical stimulation (PES), MOG1^L18F^ had greater arrhythmia incidence and inducibility (**Figure 2C-D**). In addition, mutant animals also exhibited AV conduction defects, including AV blocks (**Figure C-D**). Notably, MOG1^L18F^ mice exhibit arrhythmias at both low (5 Hz, 200 ms CL) and high (25-30 Hz, 40-33 ms CL) (**Figures 2C and 2E**) stimulation cycle lengths, consistent with the increase in the steepness of the QT interval restitution curve, a typical feature of LQT3^16^ (**Figure 2F**).

**Figure 2.**
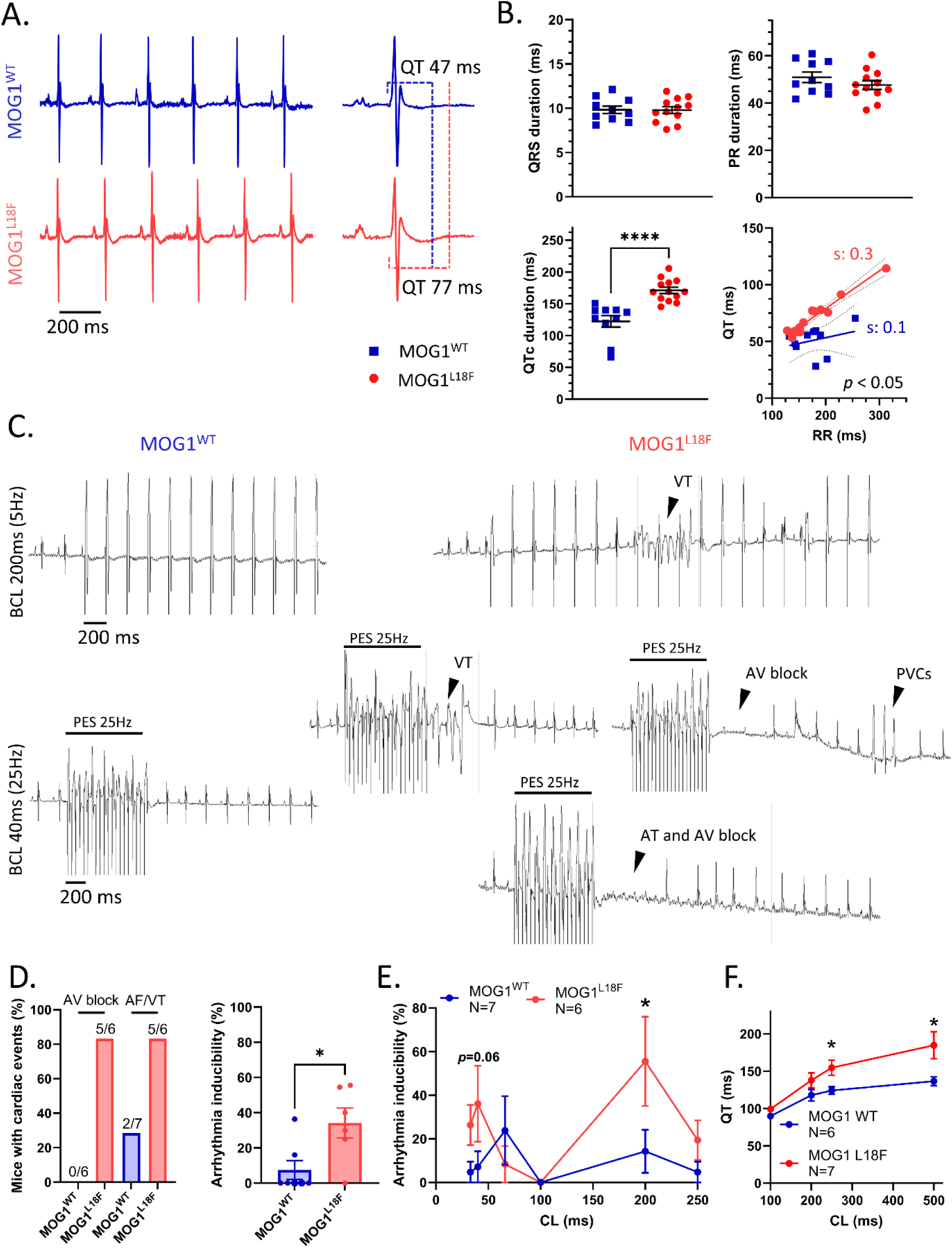
MOG1^L18F^ mice recapitulate the electrical phenotype of the patient. **A.** Representative lead-II ECG recordings in MOG1^WT^ (top, blue) and MOG1^L18F^ (bottom, red) anesthetized mice, indicating the QT interval at the right graphs. **B.** Quantification of QRS, PR, QT and QTc intervals in MOG1^WT^ and MOG1^L18F^ mice. QTc interval was prolonged in MOG1^L18F^ mice, and the slope (s) of the QT vs RR dependence is greater than in MOG1^WT^ mice. Every value represents the average of five consecutive beats. **C.** Representative surface ECG recordings during PES at 5 Hz (top) and 25 Hz (bottom) in both mice models. Different arrhythmic events are indicated with black arrows (VT, AV block, PVCs and AT). **D.** Incidence of AV block and AF/VT, and arrhythmia inducibility in MOG1^WT^ and MOG1^L18F^ mice after ventricular programmed electrical stimulation (PES) (total arrhythmia inducibility was calculated when stimulating at 4, 5, 25 and 33 Hz). **E.** Inducibility of AF/VT after pacing at different cycle lengths (CL) (from 250 to 33 ms). **F.** QT interval measured at different CL (from 500 to 200 ms). Each measure represents the average of ten beats. For every panel, each value is represented as mean±SEM. Statistical analyses were conducted using one-way ANOVA, followed by Tukey’s multiple comparisons (B), two-tailed t-test (D, left), Fisher’s exact test (D, right), Multiple unpaired t-test (E) and two-way ANOVA, followed by Sídák’s multiple comparisons (F). *P < 0.05; ****P < 0.0001.

#### Hypokalemia enhances arrhythmias in MOG1^L18F^ mouse hearts

To determine whether the MOG1^L18F^ mouse reproduced the arrhythmias observed in the patient, including nsVT, we performed optical mapping in isolated, Langendorff-perfused hearts paced at various frequencies. We tested for arrhythmia inducibility under normokalaemia (5.4 mM K^+^) and hypokalemia (2.7 mM K^+^) to mimic the condition that triggered nsVT in the patient (see **Figure 1B**). High-resolution APD maps (**Figure 3A,i, top**) and optical APs (**Figure 3A,iii, top**) of MOG1^L18F^ mouse hearts paced at 200 ms CL (**Figure 3A**) revealed APD prolongation compared to MOG1^WT^ hearts. However, the APD difference became progressively less at CLs shorter than 150, giving rise to an APD_80_ restitution curve that was steeper than that of MOG1^WT^ hearts (**Figure 3A,ii and S2A**), with a CV restitution that was less dependent on the CL (**Figure 3B,ii**). Moreover, under hypokalemia, the APD restitution of MOG1^L18F^ hearts became even steeper (**Figure 3A,ii**), while the CV velocity restitution levelled (**Figure 3B,ii**), both conditions contributing to the higher inducibility of arrhythmias observed in **Figure 2**. This effect washed out when restoring the K^+^ to 5.4 mM (**Figure S2B**).

**Figure 3.**
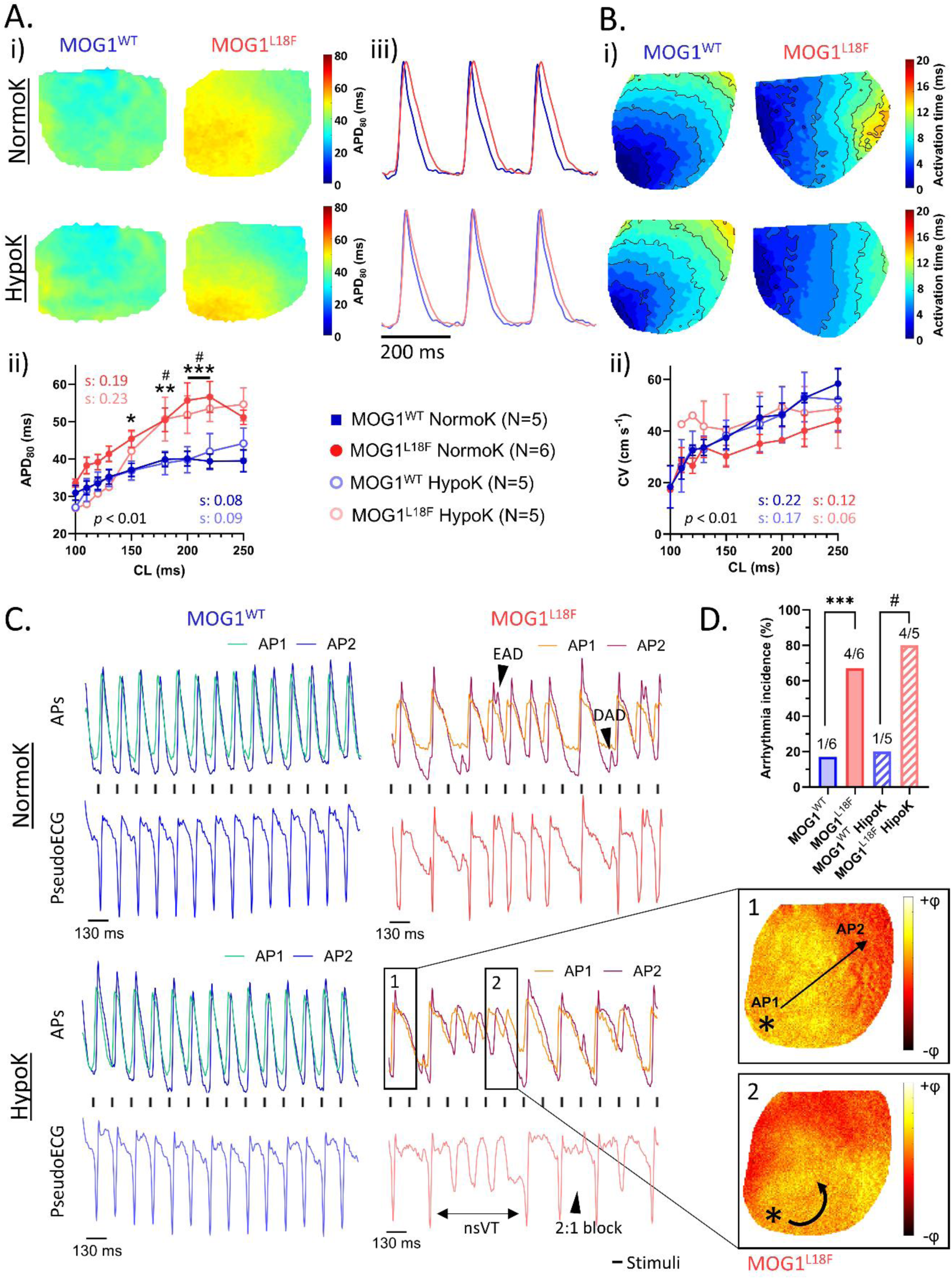
Hypokalemia worsens arrhythmia in MOG1^L18F^ mice. A. **i)** Representative APD_80_ maps from optical mapping experiments in MOG1^WT^ (left) and MOG1^L18F^ (right) hearts stimulated at 200 ms CL under normokalaemia (NormoK, up) and hypokalemia (HypoK, down). **ii)** APD_80_ restitution curve showing a slower repolarization in MOG1^L18F^ hearts specially at lower frequencies, the slope (s) is indicated. **iii)** Propagated optical action potential in MOG1^WT^ (blue) and MOG1^L18F^ (red) hearts stimulated at 200 ms CL in normokalaemia (up) and hypokalemia (down). **B. i)** Representative conduction velocity (CV) maps with 2-ms activation isochrones in MOG1^WT^ (left) and MOG1^L18F^ (right) hearts stimulated at BCL of 200 ms under normokalaemia (up) and hypokalemia (down). **ii)** CV restitution curve showing a slower conduction in MOG1^L18F^ hearts, especially at lower frequencies. **C.** Single pixel optical mapping recording from the right ventricle (AP2) and the apex (AP1), and the *pseudo*ECG obtained after subtracting AP1 from AP2 in MOG1^WT^ (left) and MOG1^L18F^ (right) hearts under normokalaemia (up) and hypokalemia (down). Right maps show representative optical maps of MOG1^L18F^ when stimulated at a 130 ms BCL during hypokalemia. Asterisk (*) indicates the site of stimulation. 1 and 2 correspond to its homonymous in the *pseudo*ECG, note the rotor in 2. **D.** Contingency plots of the number of hearts with arrhythmia in each condition. Each value is the mean±SEM (N=5-6 per condition; 2-way ANOVA corrected by the Šídák multiple comparisons test (A), and Fisher’s exact test (D); *P<0.05; **P<0.01; ***P<0.001 in NormoK; ^#^P<0.05 in HypoK).

Unlike MOG1^WT^, MOG1^L18F^ hearts exhibited a high incidence of complex conduction block patterns under both normokalaemia and hypokalaemia, as shown in **Figure 3C and D** and **Figure S3**. Single-pixel optical APs recorded from two different locations—the ventricular apex (AP1) and the right ventricular base (AP2)—revealed that some areas of the MOG^1L18F^ ventricles failed to consistently follow the stimulus cycle length (CL) at 130 ms. These regions displayed progressive conduction delays with intermittent blocks, secondary to prolonged repolarization (**Figure 3C,1**), and developed early afterdepolarizations (EADs) (**Figure 3C**). Spatiotemporal dispersion of repolarization, resulting in slow and heterogeneous conduction, is evident in the maps and single-pixel APs of **Figure 3C** and **S3**; isochronal crowding is apparent in some activation maps (**Figure S3A**). Hypokalaemia further exacerbated these phenomena (**Figure S3A,ii**), leading to more pronounced conduction alterations. **Figure S3B,ii** demonstrates conduction failure in one direction on the optically recorded ventricular surface, with retrograde conduction occurring via the unobserved surface (noting that the video camera captures only the anterior surface). This pattern recurred over several beats, resulting in an optical pseudoECG that appeared as bidirectional tachycardia. Collectively, these changes in cardiac conduction predisposed to more severe arrhythmias, such as non-sustained ventricular tachycardia (nsVT), consistent with the patient’s clinical presentation. The arrhythmogenicity was markedly increased under hypokalaemia, recapitulating the patient’s phenotype (**Figure 3D**).

#### MOG1^L18F^ increases *I*_NaL_ in mouse ventricular cardiomyocytes

Since MOG1 modulates Na_V_1.5 channels^6^, we performed patch-clamp experiments in isolated cardiomyocytes to study the effects of MOG1^L18F^ on the sodium current (*I*_Na_). On voltage-clamping, compared to that of MOG1^WT^ cardiomyocytes, peak *I*_Na_ was slightly increased (**Figure 4A, 4C**), but *I*_NaL_ was dramatically enhanced in MOG1^L18F^ (**Figure 4A-C**), which increased the *I*_NaL_ to peak *I*_Na_ ratio in the same direction (**Figure 4D**). In MOG1^L18F^ cardiomyocytes, the increase in *I*_NaL_ was more pronounced at membrane potentials close to 0 mV, and the maximum range of *I*_NaL_ (-30 to -5 mV) did not overlap with the range of peak *I*_Na_ (-45 mV), or with the maximum range of *I*_NaL_ in MOG1^WT^ cardiomyocytes (-60 to -40 mV) (**Figure 4C-D**). The voltage dependence of activation and steady-state inactivation was not modified (**Table S1**), but the Na^+^ window current recorded in MOG1^L18F^ cardiomyocytes was larger than that in MOG1^WT^, due to the incomplete inactivation of *I*_Na_, increasing its open probability (**Figure 4E**). Additionally, the recovery from inactivation was faster in MOG1^L18F^ than in MOG1^WT^ cardiomyocytes (**Figure 4F, Table S1**). RT-qPCR and total protein analysis showed similar *Scn5a* and Na_V_1.5 mRNA and protein levels (**Figure 4Gi-ii**), and membrane enrichment revealed comparable sarcolemmal expression of Na_V_1.5 (**Figure 4Giii-iv**) in MOG1^WT^ and MOG1^L18F^ cardiomyocytes. These findings demonstrated that the MOG1^L18F^ variant enhances the *I*_NaL_ without altering the effects of MOG1 on Na_V_1.5 expression or trafficking (**Figure 4G**). In addition, no changes in ventricular K^+^ currents were detected **(Figure S4)**.

**Figure 4.**
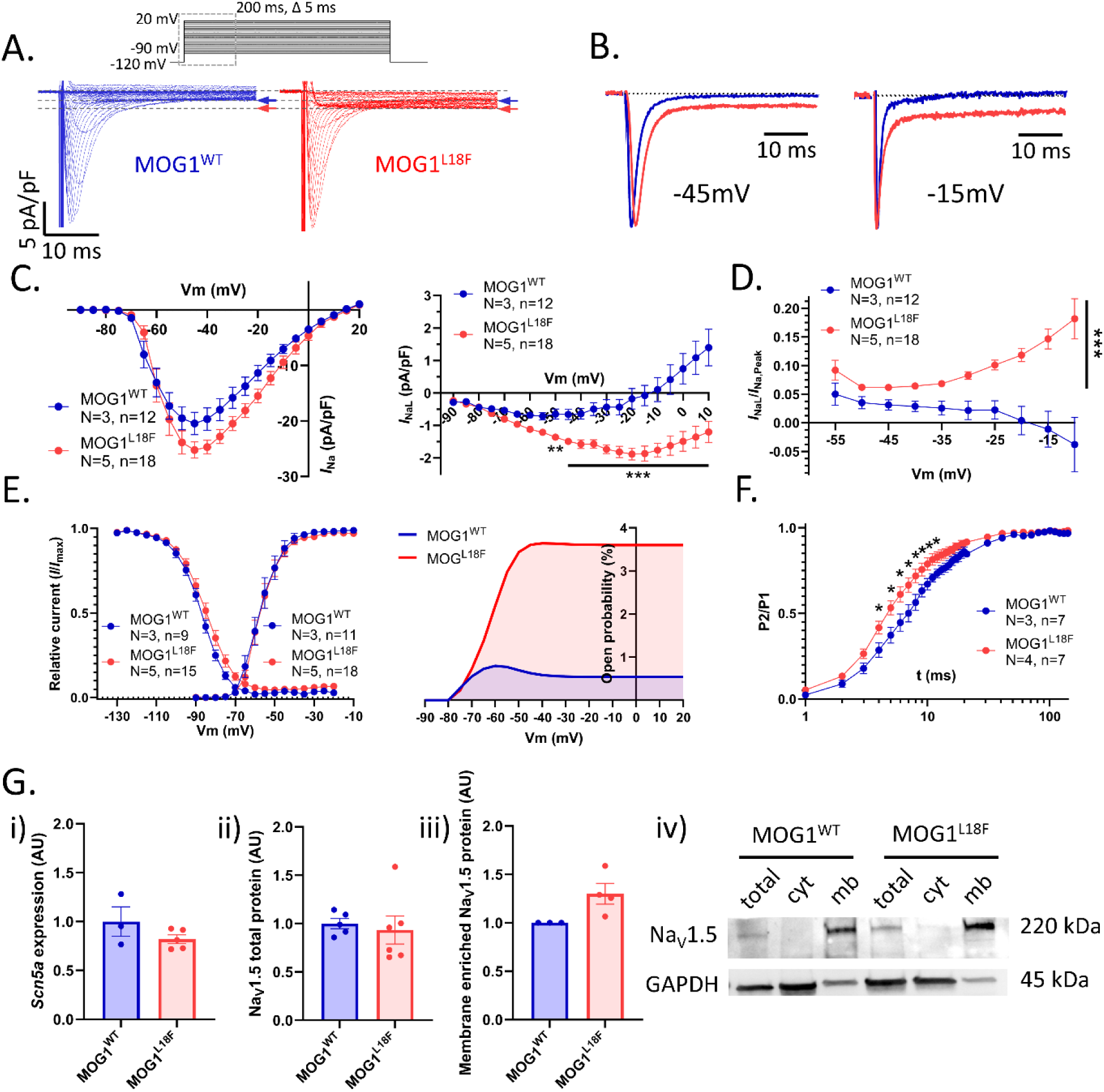
MOG1^L18F^ enhances *I*_NaL_ in mouse ventricular cardiomyocytes. **A.** Representative *I*_Na_ current traces obtained after applying the voltage-clamp protocol shown in the upper panel in MOG1^WT^ (left, blue) and MOG1^L18F^ (right, red) ventricular cardiomyocytes. Blue and red arrows indicate the *I*_NaL_ magnitude recorded from MOG1^WT^ and MOG1^L18F^ cardiomyocytes, respectively. **B.** Superimposed normalized Na^+^ currents of MOG1^WT^ (blue) and MOG1^L18F^ (red) at -45 (left) and -15 mV (right). **C.** Current-amplitude (I-V) relationship of the current density of peak (left) and late (right) Na^+^ current in both mouse models. *I*_NaL_ was enhanced in MOG1^L18F^ cardiomyocytes, especially at more depolarized membrane potentials **D.** I-V relationship of the *I*_NaL_/*I*_Na_ ratio. **E.** Activation and steady-state inactivation curves (left) and open probability of Na_V_ channels (right) calculated as the product of the fitting to a Boltzmann equation of activation and inactivation curves in each condition. **F.** Recovery from inactivation of MOG1^WT^ and MOG1^L18F^ cardiomyocytes. **G.** Quantification of *Scn5a* mRNA (**i**), Na_V_1.5 total protein levels (**ii**), as well as Na_V_1.5 protein expression in enriched membrane fraction (**iii**). Panel **iv** shows a representative immunoblot of Na_V_1.5 in the total lysate, the cytoplasm and membrane enriched fractions in both WT and L18F cardiomyocytes. Data are expressed as mean±SEM. Statistical analyses were conducted using two-way ANOVA, followed by Sídák’s multiple comparisons (C-E), or one-way ANOVA, followed by Tukey’s multiple comparisons (F-G). *P < 0.05; ***P < 0.001.

To test whether the above effects were produced directly on Na_V_1.5 channels, we transfected HEK/Na_V_1.5 cells with mCherry vector (Control), MOG1^WT^, or MOG1^L18F^ expression vectors. The results were different from those in cardiomyocytes; MOG1^L18F^ increased peak *I*_Na_ without modifying *I*_NaL_ (**Figure S5A-B**). The voltage dependence of activation and inactivation, as well as the recovery from inactivation, was not modified in MOG1^L18F^-transfected cells in comparison to those transfected with MOG1^WT^ (**Figure S5C-D, Table S2).** The differences between cardiomyocytes and HEK/Na_V_1.5 cells suggest that the effects of MOG1 ^L18F^ were not due to a direct effect on Na_V_1.5 channels, pointing to a cardiac-specific mediator in such processes.

#### MOG1^L18F^ enhances *I*_NaL_ via activation of the Na_V_1.8-short isoform (Na_V_1.8-s)

As previously mentioned, *I*_NaL_ is regulated by different proteins, such as CaMKII, and neuronal Na_V_ channels like Na_V_1.8^16,17^. To decipher the molecular mechanisms underlying the *I*_NaL_ increase revealed in MOG1^L18F^ cardiomyocytes, we tested the effects of a selective Na_V_1.8 inhibitor (A-803467) and a CaMKII inhibitor (KN-93) on MOG1^L18F^ cardiomyocytes (**Figure 5A-D**). The increased *I*_NaL_ in MOG1^L18F^ cardiomyocytes was reversed by the selective antagonist of Na_V_1.8, A-803467, to resemble MOG1^WT^ currents, but not by the CaMKII blocking with KN-93 (**Figure 5A-C**). Na_V_ channels’ open probability was also reduced by A-803467, especially at membrane potentials positive to -60 mV (**Figure 5D**). These results strongly indicate that Na_V_1.8 channels mediate the increase in *I*_NaL_. To delve into this mechanism, we first checked out *Scn10a* and Na_V_1.8 expression levels in cardiomyocytes (**Figure 5E**). By qPCR and western blot, we could detect the cardiac short isoform of *Scn10a* and Na_V_1.8 (Na_V_1.8-s) described by Man et al., 2021^23^, but not the full-length mRNA or protein (**Figure S6**), with no differences in total *Scn10a-s* and Na_V_1.8-s expression levels (**Figure 5Ei-ii,iv)**. However, membrane enrichment revealed a significantly increased of Na_V_1.8-s in MOG1^L18F^ cardiomyocytes (**Figure 5Eiii-iv**), thus endorsing the implication of Na_V_1.8-s on the phenotype.

**Figure 5.**
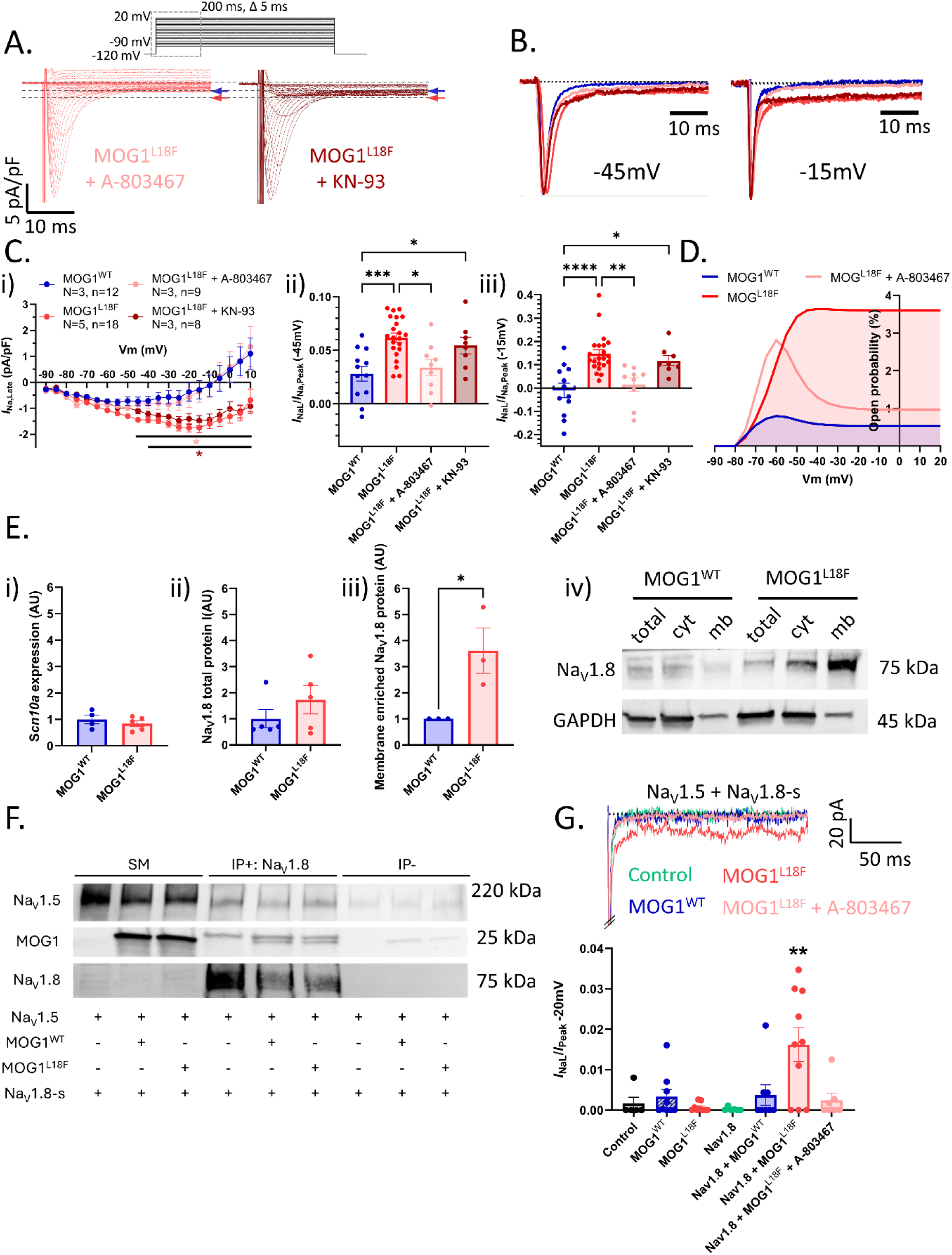
*I*_NaL_ increase is mediated through Na_V_1.8-s channels in MOG1^L18F^ mice. **A.** Original current recordings of MOG1^L18F^ ventricular cardiomyocytes treated with A-803467 (left, pink) or KN-93 (right, brown). **B.** Superimposed normalized Na^+^ currents of MOG1^WT^ (blue), MOG1^L18F^ (red) and MOG1^L18F^ + A-803467 (pink) cardiomyocytes at -45 (left) and -15 mV (right). **C.** I-V relationship of *I*_NaL_ (**i**), and the *I*_NaL_/*I*_Na_ ratio at -45 mV (**ii**) and -15 mV (**iii**) recorded from ventricular cardiomyocytes pertaining to all experimental conditions. **D.** Open probability of Na_V_ channels calculated as the product of the fitting to a Boltzmann equation of activation and inactivation curves in each condition. **E.** Quantification of *Scn10a* mRNA (**i**), Na_V_1.8 total protein levels (**ii**), as well as Na_V_1.8 protein expression in enriched membrane fraction (**iii**). Representative immunoblot of Na_V_1.8 in the total lysate, the cytoplasm and membrane enriched fractions in both WT and L18F cardiomyocytes (**iv**). **F.** HEK/Na_V_1.5 cells were cotransfected with Na_V_1.8-s with (+) or without (-) MOG1^WT^ and MOG1^L18F^. Total lysates were immunoprecipitated (IP) against Na_V_1.8 and immunoblotted (IB) against Na_V_1.5, MOG1 and P2A (Na_V_1.8) (n=3). SM: starting material, IP+: IP in the presence of antibody, IP-: IP in the absence of antibody. **G.** Representative current recording of *I*_Na_ at -20 mV in HEK/Na_V_1.5 cells transfected with Na_V_1.8-s plus mCherry, MOG1^WT^ or MOG1^L18F^, and the latter also treated with A-803467. Lower graph represents the quantification of *I*_NaL_/*I*_Na,Peak_ in all experimental conditions. Data are expressed as mean±SEM. Statistical analyses were conducted using two-way ANOVA, followed by Sídák’s multiple comparisons (C-F), or one-way ANOVA, followed by Tukey’s multiple comparisons (G). *P < 0.05; **P < 0.01; ***P < 0.001; ****P<0.0001. In Ci, pink asterisk (*) refers to MOG1^L18F^ + A-803467 vs MOG1^L18F^, and brown asterisk (*) to MOG1^L18F^ + KN-93 vs MOG1^WT^.

To further investigate the underlying molecular mechanism, we conducted coimmunoprecipitation experiments in HEK293 cells to test whether the MOG1^L18F^ variant alters the Na_V_1.5/Na_V_1.8-s/MOG1 complex. Immunoprecipitation of Na_V_1.8-s consistently pulled down Na_V_1.5 and MOG1 in co-transfected HEK/Na_V_1.5 cells (**Figure 5F**), indicating that the MOG1^L18F^ variant does not affect the level of MOG1 interaction with the Na_V_1.5/Na_V_1.8-s complex. Additional experiments demonstrated that MOG1^WT^ and MOG1^L18^ also bound to Na_V_1.8-s in the absence of Na_V_1.5 (**Figure S7A**). The similar interactions observed with MOG1^WT^ and MOG1^L18F^ suggest that the variant does not alter binding affinity or stoichiometry. Therefore, the increased *I*_NaL_ can be due to altered channel gating or regulation. To test whether the effects on *I*_NaL_ were produced by Na_V_1.8-s current, we co-transfected HEK293 cells with the cardiac isoform Na_V_1.8-s in the absence and the presence of MOG1^WT^ or MOG1^L18F^, but no current was detected (data not shown). This fact suggested that the observed effects of MOG1^L18F^ on Na_V_1.8-s, and consequently on I_NaL_, were produced by an indirect mechanism, likely by Na_V_1.8-mediated modulation of Na_V_1.5. To test this hypothesis, we co-transfected HEK/Na_V_1.5 cells with Na_V_1.8-s in the absence and the presence of MOG1^WT^ or MOG1^L18F^, recapitulating the *I*_NaL_ increase observed in MOG1^L18F^ cardiomyocytes in Na_V_1.5, Na_V_1.8-s and MOG1^L18F^ expressing cells, whilst detecting virtually no *I*_NaL_ in the other conditions (**Figure 5G**). We found no changes in the voltage-dependence of activation and inactivation (**Table S3**). As A-803467 primarily binds to S6_IV_^48^, which is present in both Na_V_1.8 isoforms, it can potentially modulate Na_V_1.8-s. Thereafter, we treated Na_V_1.5, Na_V_1.8-s and MOG1^L18F^ expressing HEK293 cells with A-803467 and reversed the *I*_NaL_/*I*_Peak_ increase **(Figure 5G)**, resembling the results obtained in ventricular cardiomyocytes **(Figure 5A-C)**. This ultimately demonstrated the selectivity of A-803467 on Na_V_1.8, its effectiveness on Na_V_1.8-s, and the implication of Na_V_1.8-s in the phenotype.

#### MOG1^L18F^ prolongs the APD and triggers arrhythmogenic activity in cardiomyocytes

On current-clamp, ventricular MOG1^L18F^ cardiomyocytes had a partially depolarized resting membrane potential (RMP) and reduced AP amplitude (**Figure 6A-B**), with no change in maximum AP upstroke velocity (dV/dt_max_), when compared to MOG1^WT^ and MOG1^L18F^ cardiomyocytes with blocked Na_V_1.8 (**Figure 6A,C**). APD at 90% repolarization (APD_90_) was dramatically increased at lower frequencies (from 0.1 to 2Hz) in MOG1^L18F^ cardiomyocytes, but was similar to MOG1^WT^ at the highest frequencies (from 4 to 5Hz) (**Figure 6D-E**). This resulted in a very steep, proarrhythmic, APD_90_ restitution curve in MOG1^L18F^ compared to MOG1^WT^ cardiomyocytes (**Figure 6D**). Moreover, the APD_70_ was also prolonged at frequencies from 0.1 to 2 Hz in MOG1^L18F^ myocytes (**Figure 6E**). However, the increased frequency-dependence of the APD in MOG1^L18F^ cardiomyocytes was reversed by A-803467 to levels similar to MOG1^WT^ cells (**Figure 6D-E**), supporting the hypothesis that MOG1^L18F^-induced cardiac electrophysiological alterations are due to its effect on Na_V_1.8 channels.

**Figure 6.**
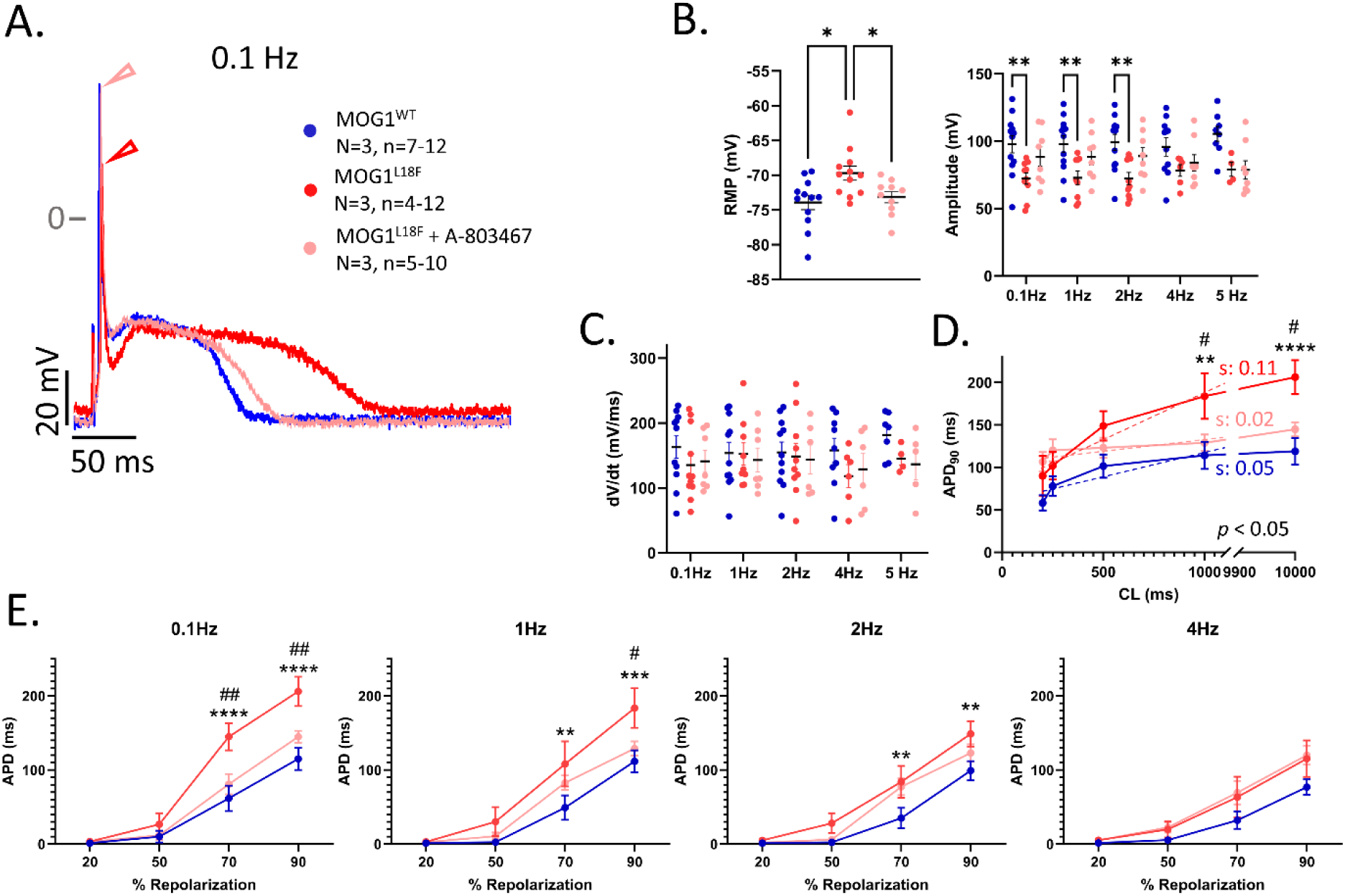
MOG1^L18F^ changes the action potential properties, being reversed by the blocking of Na_V_1.8 channels. **A.** Representative AP obtained at 0.1 Hz from MOG1^WT^ (blue), MOG1^L18F^ (red) and MOG1^L18F^ + A-803467 (pink) cardiomyocytes. **B.** Quantification of the RMP and AP Amplitude in MOG1^WT^, MOG1^L18F^, and MOG1^L18F^ treated with A-803467. **C.** Maximal upstroke velocity of the action potentials expressed as dV/dt. **D.** Restitution curve represented as the APD_90_ vs the CL between two evoked action potentials, from cycles of 1000 ms to 200 ms. Note that at higher CL, the differences between MOG1^L18F^ and the other groups are the greatest, whereas at lower CL all groups are very similar, showing differences in their slope (s). **E.** APD at different repolarizing percentages (20, 50, 70 and 90 %) and different stimulating frequencies (0.1, 1, 2 and 4 Hz) in all experimental conditions. Note that the AP morphology of MOG1^L18F^ differs more from the others at lower stimulation frequencies (between 0.1 and 2 Hz). Data are expressed as mean±SEM. Statistical analyses were conducted using one-way ANOVA, followed by Tukey’s multiple comparisons (B-C); or two-way ANOVA, followed by Sídák’s multiple comparisons (D-E). *P < 0.05; **P < 0.01; ***P < 0.001; ****P < 0.0001 when comparing MOG1^L18F^ *vs* MOG1^WT^; and ^#^P < 0.05; ^##^P < 0.01; when comparing MOG1^L18F^ vs MOG1^L18F^ + A-8033467.

As illustrated in **Figure 7**, MOG1^L18F^ cardiomyocytes exhibited more frequent spontaneous activity in the form of delayed and early afterdepolarizations (DADs and EADs, respectively), and triggered activity (TA) (92.3%, 38.5% and 61.5%, respectively), than MOG1^WT^ cardiomyocytes (35.7%, 0% and 7.1%) and MOG1^L18F^ cardiomyocytes incubated with A-803467 (35.7%, 14.3% and 7.1%) at low stimulation frequencies (**Figure 7A-B**). Additionally, unlike MOG1^WT^, some MOG1^L18F^ cardiomyocytes did not show a 1:1 (stimulus:response) ratio at frequencies higher than 4 Hz. Most importantly, the effects of the mutation on the AP properties were reversed by Na_V_1.8 inhibition with A-803467 (**Figure 7A-D**), reinforcing that the proarrhythmic phenotype of the mutant was due to a MOG1^L18F^-induced Na_V_1.8 alteration.

**Figure 7.**
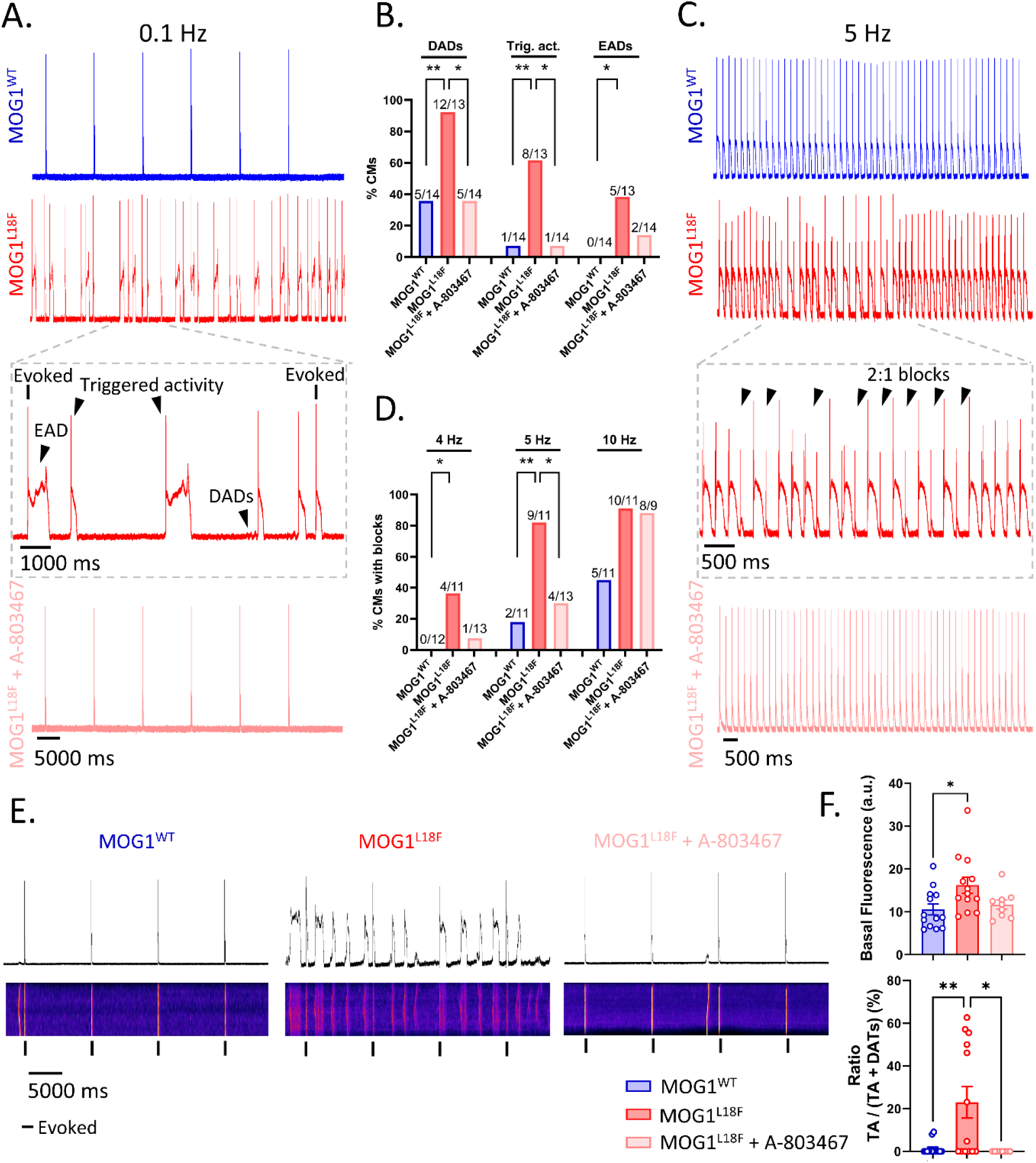
Cellular electrophysiological mechanisms underlying arrhythmia in MOG1^L18F^ mice. **A.** Representative AP traces after stimulating at 0.1 Hz MOG1^WT^ (blue), MOG1^L18F^ (red) and MOG1^L18F^ + A-803467 (pink) cardiomyocytes. Note the zoom of MOG1^L18F^ recording, highlighting some of the arrhythmic events (DADs, EADs and triggered activity). **B.** Quantification of arrhythmic activity in isolated cardiomyocytes at 0.1 Hz. **C.** Representative AP recordings of MOG1^WT^ (blue), MOG1^L18F^ (red) and MOG1^L18F^ + A-803467 (pink) cardiomyocytes stimulated at 5 Hz. In MOG1^L18F^, a zoom in which 2:1 blocks occur is shown, in which one can clearly see how in some cases the APD prolongs until the next AP cannot be triggered. **D.** Quantification of the incidence of blocks in the experimental groups at different stimulating frequencies (4, 5 and 10 Hz). **E.** Ca^2+^ dynamics and voltage recordings in response to stimulation at 0.1 Hz. Note that, in the representative MOG1^L18F^ cardiomyocyte e–c coupling shows multiple abnormal spontaneous calcium release events during both systole and diastole, which are nearly absent in MOG1^WT^ and MOG1^L18F^ + A-803467 cardiomyocytes. **F.** Quantification of the [Ca^2+^]_i_ measured as the basal fluorescence (top) and the percentage of DATs that produced triggered activity in each experimental condition (bottom). Each value is represented as mean±SEM. Statistical analyses were conducted using Fisher’s exact test (B-D) and one-way ANOVA with Tukey’s multiple comparisons (F). *P < 0.05; **P < 0.01.

Next, given the potential implications of higher Na^+^ in the intracellular Ca^2+^ homeostasis, we investigated the intracellular Ca^2+^ handling to determine whether the recorded spontaneous arrhythmic activity of MOG1^L18F^ cardiomyocytes was due to defects in calcium dynamics. As illustrated in **Figure 7E-F**, [Ca^2+^]_i_, measured as basal fluorescence, was increased in MOG1^L18F^ cardiomyocytes, as was the ratio of TA induced by delayed aftertransients (DATs) in (23.3 ± 7.4 % in MOG1^L18F^ vs 1.2 ± 0.8 % in MOG1^WT^), an arrhythmogenic alteration that was reversed by A-803467 (0%).

#### The arrhythmogenic MOG1^L18F^ phenotype was fully reversed by A-803467 *in-vivo*

To characterize the *in-vivo* implications of Na_V_1.8 in the arrhythmogenic phenotype, we treated MOG1^WT^ and MOG1^L18F^ mice with a single intraperitoneal administration of A-803467 (i.p. 20 mg/kg) or vehicle (5% DMSO, 95% PEG400). Blockade of Na_V_1.8 did not affect ECG parameters of MOG1^WT^ mice, but consistently shortened the QT and QTc of MOG1^L18F^ mice to similar values as those observed in MOG1^WT^. No changes were found in the QRS, PR and RR intervals or the P wave duration (**Figure 8A-B**). This indicates that the QT prolongation is mediated through Na_V_1.8-s, and that this channel could be a potential therapeutic target for MOG1^L18F^-induced LQTS3. Moreover, on PES, A-803467 also reduced the incidence of arrhythmia and AV block, as well as arrhythmia inducibility in MOG1^L18F^ mice, both at low and high stimulation CLs (**Figure 8C-E**), and reversed the restitution curve to values similar to those of MOG1^WT^ animals (**Figure 8F**), fully restoring normal cardiac electrical function in mutant mice. Additionally, we analyzed the effects of the sodium channel blocker flecainide, used for the treatment of LQT3^49–52^. A single i.p. administration of flecainide (40 mg/kg)^53^ increased QRS, PR, QT and RR intervals, and P wave duration in both MOG1^WT^ and MOG1^L18F^ mice, with milder effects on the former than the latter (**Figure S8A-C**). Upon intracardiac stimulation, flecainide partially decreased the number of mutant mice that presented AV block, as well as arrhythmia incidence and inducibility, but to a lesser extent than A-803467 (**Figure S8D-E**). These results suggest that a selective treatment targeting Na_V_1.8 channels could be a more promising option than a non-selective channel blocker like flecainide. This highlights the importance of understanding the molecular mechanisms of each mutation-dependent arrhythmia to determine the most appropriate treatment for patients.

**Figure 8.**
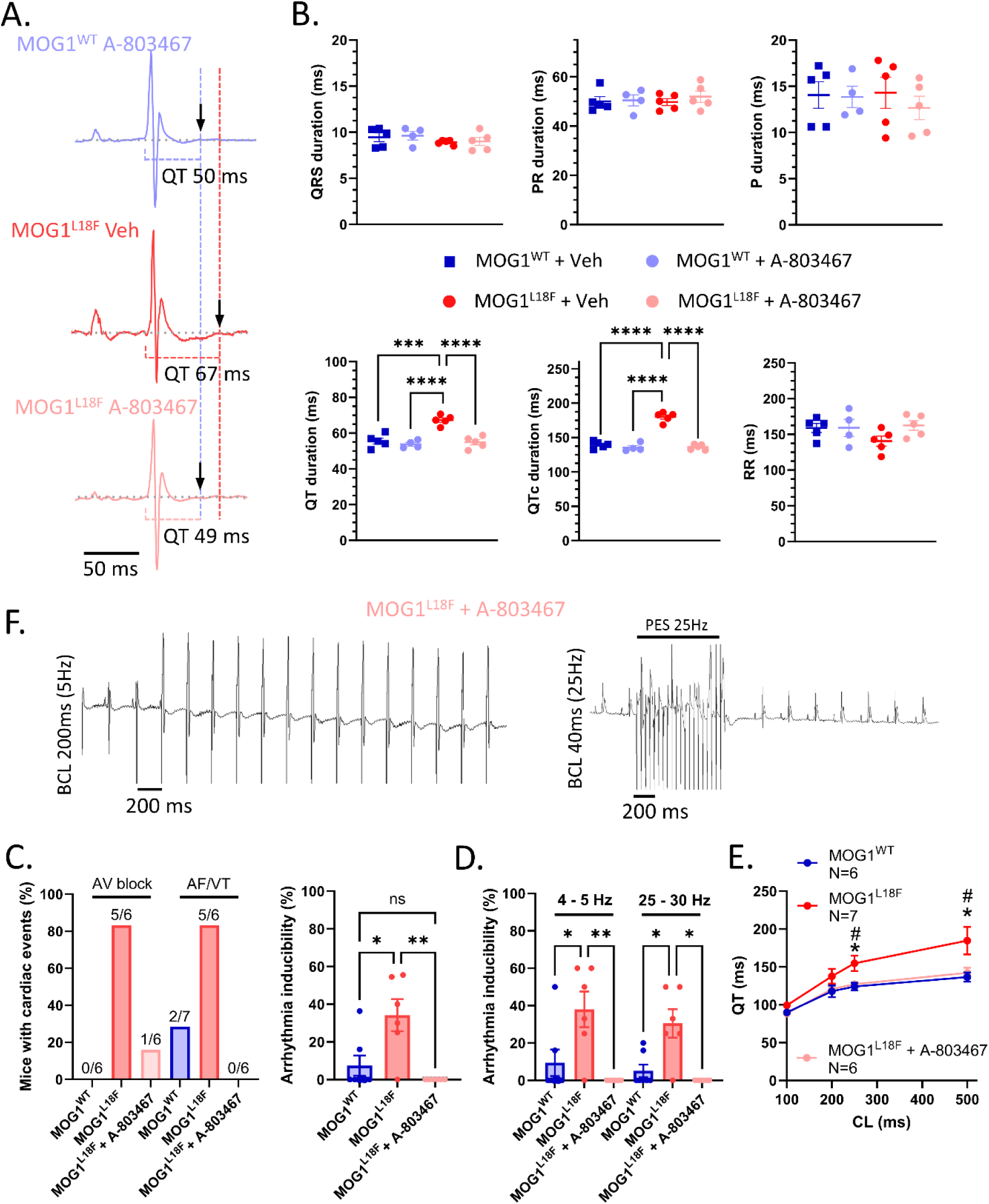
The Na_V_1.8 inhibitor A-803467 reversed the proarrhythmic electrical phenotype of MOG1^L18F^ mice. **A.** Representative lead II ECG recording of MOG1^WT^ + A-803467 (top, pale blue), MOG1^L18F^ + Vehicle (middle, red) and MOG1^L18F^ + A-803467 (bottom, pink) anesthetized mice showing the QT interval. **B.** Quantification of the ECG parameters (QRS, PR, P wave, QT, QTc and RR duration) in MOG1^WT^ and MOG1^L18F^ treated with Vehicle or A-803467. Note that the QT and QTc are the only parameters modified by A-803467 in MOG1^L18F^, but they do not change in MOG1^WT^ treated animals. **C.** Representative surface ECG recordings during PES at 5 (left) and 25 Hz (right) in MOG1^L18F^. **D.** Bar graphs showing incidence of AV block and AF/VT, and arrhythmia inducibility in MOG1^WT^, MOG1^L18F^ and MOG1^L18F^ + A-803467 mice after ventricular PES. **E.** Incidence of AF/VT after pacing at low (4-5 Hz) and high (40-33 Hz) frequencies. **F.** QT interval measured at different CL (from 500 to 200 ms). Each value is represented as mean±SEM. Statistical analyses were conducted using one-way ANOVA, followed by Tukey’s multiple comparisons (B, D right, E), Fisher’s exact test (D left); and Multiple unpaired t-test (F). *P < 0.05 when comparing MOG1^L18F^ vs MOG1^WT^, and ^#^P < 0.05 when comparing MOG1^L18F^ vs MOG1^L18F^ + A-803467.

## DISCUSSION

We characterized the functional effects of the novel MOG1^L18F^ variant, found in a patient with AV block, LQTS and polymorphic VT. We demonstrated that this variant modifies Na_V_1.8 channels increasing *I*_NaL_, which prolongs the APD and the QT interval. These findings provide robust evidence that this MOG1 variant represents a new genetic cause of LQTS.

The patient’s clinical profile closely resembled LQTS type 3 (LQT3). LQT3, caused by *gain-of-function* mutations in the *SCN5A* gene, is the third most common subtype of LQTS, accounting for approximately 5%-10% of cases^54^. On ECG, the patient exhibited QT prolongation, with longer ST segments and late-onset T waves, which is typical of LQT3^55^. Resting bradycardia, also observed in this patient, is another common finding in LQT3 patients^55^. Contrary to the more common K^+^ channel-mediated forms of LQTS^56^, in which arrhythmias are often triggered by adrenergic stress or emotional stimuli, LQT3-associated arrhythmias typically occur at rest or during periods of low activity. In addition to LQT3 features, the patient presented with 2:1 AV block, which progressed to complete AV block with AV dissociation. These conduction disturbances are typically considered distinct clinical entities. Traditionally, mutations linked to LQT3 and Brugada Syndrome (BrS) or conduction disease cause opposite biophysical effects on sodium current availability. *Gain-of-function* mutations in *SCN5A* increase *I_NaL_*, producing LQTS and atrial fibrillation. L*oss-of-function* mutations and gating or trafficking defects reduce the Na^+^ current, causing BrS and conduction defects, including AV block^14,15,57^. However, some variants produce overlapping syndromes^57^, and LQT3 patients may also present conduction disturbances, bradycardia, atrial arrhythmias, sinus node disease, or right precordial ST-segment elevation^54,58^. Upon ionic imbalance (hypokalemia), the patient exhibited polymorphic VT in the form of *Torsades de Pointes*, a malignant arrhythmia that typically appears in channelopathies like LQTS^59,60^. *Torsades de Pointes* was originally described in a LQTS patient with complete AV block^61^. The mechanism underlying this arrhythmia is a decrease in repolarization currents (K^+^ channels) or an increase in *I*_NaL_ ^62–64^. The QTc interval is usually longer than 500 ms, and VT is typically initiated after a long-coupled (450-500 ms) beat that follows a sinus or extrasystolic pause (pause-dependent or short-long-short-sequence *Torsades de pointes*)^62,64,65^, which closely resembled the abnormalities observed in our proband (**Figure 1B)**. Acute management included infusion of isoproterenol and temporary pacing. The presence of complete AV block with a low mean heart rate is an indication for permanent pacemaker implantation in children.

Genetic testing revealed that our patient had a heterozygous VUS in the *RANGRF* gene (c.52C>T), resulting in a leucine-to-phenylalanine substitution at position 18 of MOG1 (L18F). MOG1 is a cardiac-predominant regulatory protein^66^ that interacts with Na_V_1.5, enhancing its current amplitude by prompting surface expression without altering its biophysical or gating properties^6^. Notably, MOG1 does not affect other cardiac ion channels and ionic currents, suggesting specificity in modulating Na_V_1.5^11^. To date, 178 *RANGRF* variants have been reported in patients with cardiac arrhythmias on ClinVar (**Extended Table**), yet most are classified as VUS. A functional characterization is essential to classify the variants based on their causality in the disease. Also, the fact that there are so many variants in *RANGRF* found in patients with cardiac arrhythmia, but not with other diseases, might suggest a greater implication of this protein in arrhythmogenesis than that described to date. In this sense, two *loss-of-function* MOG1 variants, E83D and E61X, have been associated with BrS^7^, via reduced *I*_Na_ without changing its biophysical properties^7,8^. Additionally, due to its chaperone effects on Na_V_1.5, MOG1 has been reported to rescue *loss-of-function* mutations in Na_V_1.5 that cause sodium channelopathies, including BrS, dilated cardiomyopathy, and sick sinus syndrome^11,12^.

Recently, Ashraf *et al*. reported a pair of monozygotic twins with hypertrophic cardiomyopathy harboring the same MOG1^L18F^ variant found in our patient, but no functional characterization was performed^10^. Supporting a potential pathogenic role, *in vivo* zebrafish studies have implicated MOG1 in cardiac hypertrophy and heart failure^67,68^. MOG1 knock-out (KO) mice presented left ventricular (LV) systolic dysfunction associated with an increase in ventricular fibrosis, exhibiting an increased risk of arrhythmias and SCD when treated with isoprenaline^69^. Furthermore, *SCN10A* mutations have been identified in patients with LQTS^28^ and hypertrophic cardiomyopathy^29^, and Na_V_1.8 expression is increased in LV hypertrophy and heart failure in humans, enhancing the proarrhythmic *I*_NaL_^31^. Therefore, while we did not observe any structural changes in our young AAV-mediated mouse models expressing MOG1^L18F^ (animals between 20 and 30 weeks), the cellular electrophysiological effects may be compatible with those in hypertrophic cardiomyopathy. However, to our knowledge, this is the first report suggesting a potential association between MOG1 and LQTS. While traditionally considered a monogenic disease, increasing evidence points to a more complex genetic basis underlying LQTS, which may explain its variable penetrance^70,71^. Pathogenic variants in monogenic LQTS have a MAF < 0.0004%^70^. More frequent but still rare variants (MAF >0.0004 to <1%), known as LQTS-Lite-causative rare variants or functional risk alleles, can moderately impair repolarization reserve and trigger arrhythmic events only under concurrence with additional endogenous/exogenous risk factors ^70^. MOG1^L18F^, whose MAF is 0.0006%, may fall within this category – potentially explaining why this variant had not been related to LQTS before, and why our patient presented VT only upon ionic imbalance.

Our MOG^L18F^ mouse model recapitulated the patient’s phenotype, exhibiting prolonged QT intervals that are more dependent on the heart frequency, increased arrhythmia inducibility, and AV block. Optical mapping revealed that under normokalaemia, MOG1^L18F^ hearts had a prolonged APD_80_, especially at low heart frequencies, contributing to conduction block. The steeper APD_80_ restitution curve caused self-amplifying oscillations in repolarization^72^, producing APD alternants and repolarization heterogeneity -key substrates for reentrant arrhythmias, as illustrated in **Figure 3C and S3**. The CV is slightly reduced at low heart rates, which predispose the heart to reentrant circuits^73^. The arrhythmogenic phenotype of MOG1^L18F^ mice was exacerbated under hypokalemia (2.7 mM K^+^), resembling the proband’s phenotype. The APD restitution curve is even steeper under hypokalemia, thus explaining its more malignant arrhythmias. The optical *pseudo*ECG detected alterations that closely resembled those seen in the patient. Moreover, even though during sinus rhythm, overall epicardial ventricular CV was similar in MOG1^L18F^ and MOG1^WT^ hearts, the latter developed intermittent conduction defects secondary to abnormally prolonged repolarization, particularly in hypokalemia, which contributed to arrhythmia malignancy. In the patient, the QT interval was longer in normokalaemia than in hypokalemia, a phenomenon reproduced in MOG1^L18F^ but not in MOG1^WT^ hearts, thus underscoring the implication of the variant in the phenotype.

Similar to *SCN5A* LQT3 variants^57,74^, MOG1^L18F^ increased *I*_NaL_, enhancing the Na_V_1.5 channel open probability and producing a persistent inward current. Additionally, recovery from inactivation was faster. MOG1^L18F^ does not alter K^+^ currents even though Na_V_1.5 functionally and physically interacts with K_V_4.3^75^ and K_ir_2.1^76^. Importantly, HEK/Na_V_1.5 cells transfected only with MOG1^L18F^ showed no differences in *I*_NaL_, nor in the biophysical properties of the channels, compared with cells transfected with empty vector or MOG1^WT^, demonstrating that the mechanism required a mediator present in cardiomyocytes. The neural, tetrodotoxin-resistant Na_V_1.8 channel, previously linked with LQTS^28^, seemed a good candidate. This channel has several unique biophysical properties when compared to Na_V_1.5 and other Na_V_ channels, such as activation and inactivation at more depolarized voltages and slower inactivation with persistent current^21,48^. In some cardiac conditions, CaMKII activation pathologically increases *I*_NaL_ via phosphorylation of Na_V_1.5^77,78^ and Na_V_1.8^32^, but the blockade of CaMKII with the selective inhibitor KN-93, failed to produce differences between treated and non-treated MOG1^L18F^ cardiomyocytes. However, the selective Na_V_1.8 blocker, A-803467, fully abrogated the *I*_NaL_ increase and its voltage dependence shift, producing a Na^+^ current with the same characteristics as in MOG1^WT^ cardiomyocytes.

The mechanisms underlying the involvement of Na_V_1.8 in the MOG1^L18F^-induced increased *I*_NaL_ in cardiomyocytes could be explained by: 1) a greater expression of Na_V_1.8; 2) alterations in its trafficking; 3) changes in its biophysical properties; 4) modifications in the biophysical properties of Na_V_1.5 induced by Na_V_1.8; and/or 5) changes in the interactions between Na_V_1.5, Na_V_1.8 and MOG1. Further, as MOG1 modulates the expression of the transcription factor Tbx5 in zebrafish, which controls the expression of *Scn5a* and *Scn10a*, among other genes^67^, the possibility that the observed effects could be due to a transcriptional modulation was feasible, but no changes in *Scn5a* and *Scn10a* mRNAs were detected in mice, nor in Na_V_1.5 or Na_V_1.8 protein expression. Moreover, plasmalemmal expression of Na_V_1.5 was not modified by MOG1^L18F^, which indicates that this variant maintains its trafficking effects on Na_V_1.5, as shown also by the patch-clamp experiments. However, MOG1^L18F^ induced a dramatic enhancement of Na_V_1.8 membrane expression in cardiomyocytes, which could be implicated in the *I*_NaL_ enhancement. We were able to detect only the short isoform of Na_V_1.8 (Na_V_1.8-s) described by Man *et al*., 2021, instead of the full-length Na_V_1.8, both by qPCR and western blot. When transfecting Na_V_1.8-s in HEK/Na_V_1.5 cells, we obtained similar results to Man *et al*., 2021 (**Figure S7C-D**), which were close to those with the full-length Na_V_1.8^23^. This isoform is unable to conduct ions in HEK293 when transfected alone, as previously described^23^, and neither in the presence of MOG1^WT^ or MOG1^L18F^. Nevertheless, we were able to reproduce the effects of MOG1^L18F^ on *I*_NaL_ in cardiomyocytes when co-transfecting Na_V_1.8-s and MOG1^L18F^ in HEK/Na_V_1.5 cells (that is, with the three interactors expressed), and those effects were reversed with A-803467. As A-803467 binds to the pore region of III and IV domains in Na_V_1.8^48^, which is common between the short and long isoforms, this blocker can theoretically inhibit both, as confirmed by our results.

Moreover, since Na_V_1.5 and Na_V_1.8 physically interact and co-regulate^25^, and MOG1 interacts with both ion channels, we cannot distinguish whether MOG1^L18F^ exerted its effects directly on Na_V_1.8 current or indirectly, on Na_V_1.5 current via its modulation by Na_V_1.8. We demonstrated that Na_V_1.5-Na_V_1.8 interaction was not affected by MOG1^L18F^. Probably, the increase of Na_V_1.8 in the plasma membrane of MOG1^L18F^ cardiomyocytes makes the interaction between Na_V_1.5 and Na_V_1.8 more probable, as they would be located in the same cell structure. There, as they interact through their respective C-terminal (C-t) domains, Na_V_1.5 N-type inactivation might be hindered by the Na_V_1.8 C-t domain. Another complementary possibility is that the phenylalanine introduced in the 18^th^ amino acid in MOG1 produces a slight conformational change due to its aromatic nature that does not alter its interaction with Na_V_1.5 and Na_V_1.8, but hinders the introduction of the C-t of Na_V_1.5, or somehow favors that the C-t of Na_V_1.8 introduce on Na_V_1.5 inactivation gate, instead of that of Na_V_1.5, thus producing partially inactivated current, but deciphering this is beyond the scope of this study.

Several studies have demonstrated that the enhanced *I*_NaL_ initiates and maintains arrhythmias through various electrophysiological mechanisms, including EADs and DADs, automaticity, and reentry^16^. In our current clamp experiments, MOG1^L18F^ slightly depolarized the RMP, likely due to the higher amount of Na^+^ that activates the reverse function of the Na^+^/Ca^2+^ exchanger (NCX), thereby increasing [Ca^2+^]_i_. This depolarization can reduce AP amplitude, which may increase the likelihood of afterdepolarizations and triggered activity, consistent with our experiments and prior findings^16,79^. Remarkably, A-803467 fully reversed the above changes. *I*_NaL_ augmentation also reduced the repolarization reserve, prolonging the APD and the QT interval. We observed a steep increase in the APD_90_ restitution slope in MOG1^L18F^ cardiomyocytes, which indicates a greater rate dependence of repolarization compared to MOG1^WT^ or MOG1^L18F^ treated with A-803467. A steep APD restitution slope is a recognized mechanism for dynamic instability, which can lead to APD alternants formation^80^, as confirmed in our optical mapping experiments. Several mechanisms may explain the increased restitution slope: Na^+^ accumulation or incomplete recovery of Na_V_ channels at higher pacing rates, and the reverse-use dependence of *I* ^16^. These effects generate spatiotemporal dispersion of repolarization, generating gradients of refractoriness that may promote local conduction block and re-entrant substrates, predisposing to *Torsades de Pointes* and ventricular fibrillation^81,82^, as observed in the optical mapping experiments (see **Figure 3**). The prolonged APD also facilitated the formation of EADs, while altered Ca^2+^ handling led to DADs and triggered activity. Compared to MOG1^WT^, a higher proportion of delayed aftertransients (DATs) in MOG1^L18F^ cardiomyocytes led to DADs that triggered APs to generate premature ventricular excitations^16,83^. These events were suppressed by inhibiting *I*_NaL_ by A-803467 in MOG1^L18F^ cardiomyocytes, reinforcing the role of Na_V_1.8 in altering Na^+^ and, consequently, Ca^2+^ dynamics in the mutant cardiomyocytes. Ultimately, elevated *I*_NaL_ creates both a substrate (spatiotemporal dispersion of repolarization and refractoriness) and triggers (EADs, DADs, and triggered activity), which together increase the risk of malignant arrhythmias^16^ – consistent with the clinical phenotype of the patient and the mouse harboring MOG1^L18F^ variant.

Additionally, these cellular electrophysiological changes can explain the effects of hypokalemia on MOG1^L18F^ hearts. MOG1^L18F^ cardiomyocytes have increased *I*_NaL_, activating NCX in the reverse mode, thus elevating [Ca^2+^]_i_, causing partial depolarization. When lowering extracellular K^+^, even though *I*_K1_ current is reduced, which destabilizes the RMP, the K^+^ equilibrium (E_K_) is shifted towards more negative potentials, thus hyperpolarizing the cardiomyocytes^84^. The hyperpolarized RMP increases fast Na^+^ channel availability, enhancing peak *I*_Na_ and CV. At the same time, the reduced Na^+^ driving force during the *plateau* diminishes *I*_NaL_, favoring a faster repolarization, especially at higher stimulation frequencies ^84^. Thus, the AP is shortened at higher stimulation frequencies, increasing the steepness of the restitution curve, which enhances the heterogeneity of repolarization and prompts arrhythmia.

Treatment with A-803467 (20 mg/kg) abridged the QT interval prolongation, improved AV conduction and reduced arrhythmia susceptibility in MOG1^L18F^ mice. A-803467 also normalized the steep restitution slope, effectively reversing the proarrhythmic phenotype, and demonstrating that Na_V_1.8 dysfunction induced by the MOG1^L18F^ variant underlies the observed electrical abnormalities in MOG1^L18F^ mice. While sodium channel blockers, including mexiletine^85^, flecainide^49–52^ and ranolazine^86^ have been employed for LQT3 treatment, our data showed that flecainide was less effective than A-803467 when treating MOG1^L18F^. In addition, in both MOG1^WT^ and MOG1^L18F^ mice, flecainide produced alterations in the ECG parameters that could lead to arrhythmia. In our experiments, A-803467 (20 mg/kg) did not modify any ECG parameters in WT animals, differing from prior studies using higher doses (100 mg/kg)^87^, where QRS and PR intervals were prolonged, likely due to blockade of peak *I*_Na_. Given that A-803467 selectively inhibits Na_V_1.8 at low concentrations (IC_50_ ∼ 0.03 µM) without significantly blocking peak *I*_Na_ (> 3 µM)^88^, we selected a lower dose based on analgesic studies to minimize the off-targets on normal nociception or on Na_V_1.5 channels^89,90^. Moreover, A-803467 did not impair motor function or coordination at doses up to 300 mg/kg, i.p. in mice or rats^90^. Considering A-803467’s pharmacokinetic properties^91^, future research should explore alternative Nav1.8 inhibitors with a better pharmacokinetic profile; for example, VX-548, which has recently completed phase III clinical trials for acute pain^92,93^. In the future, this compound might be repurposed for LQTS in which Na_V_1.8 is augmented.

### Limitations

Certain potential limitations of our study should be considered. First, our work is based on a mutation discovered in a single patient with no family history of cardiac disease. However, the ECG findings of QTc prolongation, together with the favorable genotype of a *RANGRF* (MOG1) mutation, strongly support the relevance of our study. In addition, QT interval assessment in this patient was only possible in the presence of AV block. Second, we are well aware of the limitations of using mouse models to study cardiac arrhythmia mechanisms associated with human channelopathies. Mice lack many of the ion channels and electrophysiological properties that characterize human AP, and any comparison or extrapolation must be made with caution. Third, as Na_V_1.5 and Na_V_1.8-s physically interact and co-regulate^25^, we could not establish whether MOG1^L18F^ exerted its effects directly on Na_V_1.8 current or indirectly on Na_V_1.5 current via its modulation by Na_V_1.8-s. Full-length Na_V_1.8 does not produce detectable functional currents when transiently transfected into non-neuronal cell lines, with the C-t of the protein identified as the limiting factor^94^. However, maybe its interaction with Na_V_1.5 enables its conductance. In contrast, Na_V_1.8 does generate functional currents in neuronal cell lines, but these cells contain abundant endogenous Na_V_ channels, which introduce background noise into the results. Although our cardiomyocyte observations were reproduced in HEK293 cells, further work is required to dissect the precise molecular interplay between MOG1^L18F^, Na_V_1.5 and Na_V_1.8. Lastly, despite some controversial results about whether full-length Na_V_1.8 is expressed in cardiomyocytes, some groups have detected it in human cardiomyocytes^31,33^. Therefore, we cannot rule out the possibility that the full-length isoform is below the threshold for detection in cardiomyocytes under our experimental conditions. Addressing each and all the above mechanisms is well beyond the scope of the present study and will require additional work. Despite these constraints, our results unambiguously demonstrate the relevant contribution of Na_V_1.8 to the proarrhythmic *I*_NaL_ and QT prolongation in our model.

## Conclusion

We demonstrate, for the first time, that cardiac-specific expression of MOG1^L18F^ in mice impairs AV conduction and increases *I*_NaL_, which prolongs the APD and the QT interval, recapitulating the patient’s arrhythmogenic LQTS phenotype. We further identify Na_V_1.8-s as the principal contributor to the increase in *I*_Na_, highlighting a therapeutic target for LQTS. Hence, this is the first demonstration that a variant in MOG1 increases *I*_NaL_, providing a new genetic basis for LQTS.

## Data Availability

All data produced in the present study are available upon reasonable request to the authors

## Acknowledgements

The authors acknowledge Dr. Huges Abriel (University of Bern, Bern, Switzerland) for kindly providing us with the HEK/Na_V_1.5 cells, and Dr. Phil Barnett (Amsterdam UMC, University of Amsterdam) for providing the Na_V_1.8-s vector. We thank Dr. Eva Cabrera-Borrego, Dr. Patricia Sánchez-Pérez and Dr. Amaia Talavera-Gutiérrez for providing discussions and revisions. The authors thank the Centro Nacional de Investigaciones Cardiovasculares (CNIC) Viral Vectors Unit for producing the AAV9. We are grateful to the patient and family members for agreeing to participate in this study.

## Funding

Supported by Grant PID2022-137214OB-C21; funded by MCIN/AEI/10.13039/501100011033; Grant CB/11/00222 funded by Instituto de Salud Carlos III CIBERCV (CB/11/00222); Grant S2022/BMD-7223 by Comunidad Autónoma de Madrid (CAM) (to C.V.); Grants FPU17/02731 (to P.G.S.), FPU22/03253 (to J.M.R.R) and FPU20/01569 (to A.I.M.M); funded by Ministerio de Ciencia e Innovación. Supported also by National Heart, Lung and Blood Institute, NIH, grant number R01HL163943; La Caixa Banking Foundation project code HR18-00304 (LCF/PR/HR19/52160013); grants PI-FIS-2020 # PI20/01220 and PI-FIS-2023 # PI23/01039 from Instituto de Salud Carlos III (ISCIII) and co-funded by Fondo Europeo de Desarrollo Regional (FEDER), and by The European Union, respectively; grant PID2020-116935RB-I00 and BFU2016-75144-R funded by MCIN/AEI/10.13039/501100011033; Fundación La Marató de TV3 (736/C/2020) “amb el suport de la Fundació La Marató de TV3”; European Union’s Horizon 2020 grant agreement GA-965286; and Program S2022/BMD7229 -CM ARCADIA-CM funded by Comunidad de Madrid (to JJ).

## Author Contributions

P.G.S., C.V. and J.J. conceived the study and co-designed the experiments; P.G.S., A.M. and A.V-Z. carried out cellular electrophysiology experiments; F.M.C. generated the mouse models; P.G.S. and F.M.C. performed the *in-vivo* characterization; J.M.R performed the optical mapping experiments; A.M. carried out the intracellular Ca^2+^ dynamics experiments. A.J.C. provided the clinical data; C.P. performed the genetic analysis of the patient; A.I.M.M. provided technical support, discussions, and revisions; P.G.S. and J.J. co-wrote the manuscript; C.V. and J.J. provided supervision, funding, and revisions. All authors discussed the results, commented on the manuscript, and approved it.

## Conflict of interest

The authors declare no competing interests.

## Notes

### Competing Interest Statement

The authors have declared no competing interest.

### Author Declarations

Informed consent from the parents was obtained for the use of clinical data with scientific objectives in the Hospital Universitario La Paz of Madrid, Spain. Ethical oversight was waived by the Comite de etica de la investigacion con medicamentos (CEIM) from Hospital Universitario La Paz.

